# High prevalence of symptoms among Brazilian subjects with antibodies against SARS-CoV-2: a nationwide household survey

**DOI:** 10.1101/2020.08.10.20171942

**Authors:** Ana M B Menezes, Cesar G Victora, Fernando P Hartwig, Mariângela F Silveira, Bernardo L Horta, Aluísio J D Barros, Marilia A Mesenburg, Fernando C Whermeister, Lúcia C Pellanda, Odir A Dellagostin, Cláudio José Struchiner, Marcelo N Burattini, Fernando C Barros, Pedro C Hallal

## Abstract

Since the beginning of the pandemic of COVID-19, there has been a widespread assumption that most infected persons are asymptomatic. A frequently-cited early study from China suggested that 86% of all infections were undocumented, which was used as indirect evidence that patients were asymptomatic.

Using data from the most recent wave of the EPICOVID19 study, a nationwide household-based survey including 133 cities from all states of Brazil, we estimated the proportion of people with and without antibodies for SARS-CoV-2 who were asymptomatic, which symptoms were most frequently reported, the number of symptoms reported and the association between symptomatology and socio-demographic characteristics. We were able to test 33,205 subjects using a rapid antibody test that was previously validated. Information on symptoms was collected before participants received the test result. Out of 849 (2.7%) participants who tested positive for SARS-CoV-2 antibodies, only 12.1% (95%CI 10.1-14.5) reported no symptoms since the start of the pandemic, compared to 42.2% (95%CI 41.7-42.8) among those who tested negative. The largest difference between the two groups was observed for changes in smell or taste (56.5% versus 9.1%, a 6.2-fold difference). Symptoms change in smell or taste, fever and myalgia were most likely to predict positive test results as suggested by recursive partitioning tree analysis.

Among individuals without any of these three symptoms (74.2% of the sample), only 0.8% tested positive, compared to 18.3% of those with both fever and changes in smell or taste. Most subjects with antibodies against SARS-CoV-2 in Brazil are symptomatic, even though most present only mild symptoms.

## INTRODUCTION

Since the beginning of the pandemic of COVID-19, there is a widespread notion that most people infected by SARS-CoV-2 are asymptomatic, following an early article from China stating that 86% of those infected did not report any symptoms.^1^ More recently, several clinical studies became available, showing that the prevalence of asymptomatic infected individuals ranges from 4% to 75%.^2-6^ These discrepancies might be explained by the use of different lists of symptoms, different recall periods, as well as different populations. Population-based studies are particularly relevant for studying SARS-CoV-2 symptoms, because asymptomatic patients or those with mild symptoms may be identified at home, rather than in health service-based studies.

Using data from the most recent wave of the EPICOVID19 study, a nationwide household-based survey including 133 cities from all states of Brazil,^7^ we estimate the proportion of people with and without antibodies for SARS-CoV-2 who were asymptomatic. We investigated which symptoms were most frequently reported, how many symptoms were reported by each subject, and the associations between symptoms and sociodemographic characteristics. We also performed conditional inference tree analyses using binary recursive partitioning to identify which combinations of symptoms were most likely to predict positive test results.

## METHODS

EPICOVID19 is a nationwide seroprevalence survey conducted in sentinel cities in 26 Brazilian states and the Federal District. The Brazilian Institute of Geography and Statistics (IBGE) divides the country into 133 intermediate regions, and the most populous municipality in each region was included in the sample. So far, the study has entailed three waves of data collection (May 14-21, June 4-7, and June 21-24). Here we report on findings from the third wave of data collection which included a detailed investigation of symptoms.

A multi-stage probabilistic sample was adopted, with 25 census tracts selected in each one of the 133 sentinel cities, with probability proportionate to size. In each sampled tract, 10 households were systematically selected, totaling 250 households per municipality. All household residents were listed, and age and sex recorded on a list. One individual was then randomly selected as the respondent for that household. Then, a finger prick blood sample was obtained and a questionnaire applied. If the selected subject did not accept to participate, a second resident was randomly chosen. In case of another refusal, the interviewers moved to the next household to the right of the one that had been originally selected. The total planned sample size was 33,250 individuals.

The WONDFO SARS-CoV-2 Antibody Test (Wondfo Biotech Co., Guangzhou, China) was used for the detection of antibodies for SARS-CoV-2 (https://en.wondfo.com.cn/product/wondfo-sars-cov-2-antibody-test-lateral-flow-method-2/); this rapid point-of-care test is based on the principle of immune assay of lateral flow and detects IgG/IgM antibodies against SARS-CoV-2. The presence of antibodies is detected by two drops of blood from a pinprick sample; after the introduction of the blood sample, valid tests are identified by a positive control line in the kit’s window; if this control line is not visible, the test is considered inconclusive. A second line also appears in the window if SARS-CoV-2-reactive antibodies are present; in the absence of antibodies, this line is not visible. This rapid test underwent independent validation studies; by pooling the results from the four validation studies, weighted by sample sizes, sensitivity was estimated at 84.8% (95% CI 81.4%;87.8%) and specificity at 99.95% (95% CI 97.8%;99.7%).^8-10^

Field workers used tablets to record the full interviews, registered all answers, and photographed the test results. All positive or inconclusive tests were read by a second observer, as well as 20% of the negative tests. Subjects were asked about presence (yes/no) of 11 symptoms since March 2020, when the first cases were reported in Brazil: fever, sore throat, cough, difficulty breathing, palpitation, change in smell or taste, diarrhea, vomiting, myalgia, shivering and headache. Subjects were classified as “asymptomatic” if they answered “no” for all symptoms.

Sociodemographic variables were also investigated: sex, age in years, schooling (last year completed/grade; recoded as primary or less; secondary; university or higher), self-reported skin color, and household assets. The official Brazilian classification of ethnicity recognizes five groups, based on the question: “What is your race or color?” The five response options are “white”, “brown” (“pardo” in Portuguese), “black”, “yellow” and “indigenous”. Interviewers were instructed to check the “yellow” option when the respondent mentions being of Asian descent, and “indigenous” when any of the multiple first nations were mentioned.^11^

The wealth index was created based on a list of assets and goods (computer or laptop, internet access, color television, air conditioning equipment, number of vehicles, cable TV, number of bathrooms and number of bedrooms), through a principal component analysis. The first component was extracted and the total sample divided into quintiles weighted by municipality urban population; the first quintile represent the 20% poorest individuals, and the fifth quintile represents the wealthiest 20% in the sample.^12^ For the schooling analysis, subjects under 5 years were excluded as they could still be attending school.

Interviewers were tested prior to the field work and only those found to be negative for the virus could participate in the study. Biological safety measures were taken to protect the health of the field workers and individual protection equipment was discarded after visiting each household. Ethical approval was provided by the Brazilian’s National Ethics Committee (process number: 30721520.7.1001.5313). Study participants were informed about the objectives of the study, possible risks and advantages. Blood collection took place after obtaining written informed consent from participants or their legal guardians. Individuals testing positive were referred to the statewide COVID-19 surveillance system. In case of a positive rapid test by the respondent, all other residents of the household were also tested for antibodies.

The prevalence of each of the 11 symptoms was calculated separately for individuals who tested positive and those with negative results. Means and standard errors (SE) were estimated for the variable on number of symptoms. Prevalence ratio and 95% confidence interval (95% CI) were calculated for each symptom, by dividing the frequency of each symptom in positive and negative subjects. Chi-squared test for heterogeneity or linear trend were calculated, according to the type of variable studied, and interactions with the test result were also tested. Subjects with previous diagnosis of COVID-19 (n=242) and missing information on symptoms (n=1,104) were excluded from the analysis.

We also performed conditional inference tree analyses using binary recursive partitioning, accounting for multiple testing^13^. The objective of these analyses was to identify which combinations of the 11 symptoms were most likely to predict positive test results.

Analyses were performed using the software Stata version 14.1 (StataCorp, College Station, TX, USA) and conditional inference tree analyses were performed using R 3.6.1 (https://www.r-project.org/). Data will become publicly available 30 days after completion of the fieldwork at http://www.epicovid19brasil.org/.

## RESULTS

Of the target sample size comprising 33,250 individuals, we were able to include 33,205 (99.9%) participants in the study (missing information for 45 subjects). To achieve this number, a total of 59,724 houses were contacted, with 19.8% of refusals and 24.6% of houses being empty at the time of the visit. Of the 31,869 participants included (after excluding for missing on symptoms and previous COVI-19 diagnosis), 849 subjects (2.7%) tested positive for SARS-CoV-2 antibodies. Test results were only disclosed after the interview on symptoms had been completed. Table 1 shows the distribution of the sample according to sociodemographic characteristics.

**Table 1.** Distribution of the study sample according to sociodemographic characteristics and region. The EPICOVID19 study, third wave.

|  | Sample distribution |  |
| --- | --- | --- |
|  | Number | % |
| **Region** |  |  |
| Northeast | 9982 | 31.3% |
| North | 5180 | 16.3% |
| Central-West | 3603 | 11.3% |
| Southeast | 8021 | 25.1% |
| South | 5083 | 16.0% |
| **Sex** |  |  |
| Female | 18646 | 58.5% |
| Male | 13223 | 41.5% |
| **Age (years)** |  |  |
| 0-4 | 637 | 2.0% |
| 5-9 | 862 | 2.7% |
| 10-19 | 2789 | 8.8% |
| 20-29 | 4965 | 15.6% |
| 30-39 | 4999 | 15.7% |
| 40-49 | 5078 | 15.9% |
| 50-59 | 5032 | 15.8% |
| 60-69 | 4234 | 13.3% |
| 70+ | 3273 | 10.3% |
| **Color/ethnicity** |  |  |
| White | 11442 | 36.7% |
| Brown | 14131 | 45.4% |
| Black | 4264 | 13.7% |
| Asian | 897 | 2.9% |
| Indigenous | 429 | 1.4% |
| **Schooling** |  |  |
| Primary or less | 11417 | 39.3% |
| Secondary | 11363 | 39.1% |
| University or higher | 6275 | 21.6% |
| **Wealth quintiles** |  |  |
| Poorest | 7668 | 24.1% |
| 2nd | 5809 | 18.2% |
| 3rd | 6334 | 19.9% |
| 4th | 6214 | 19.5% |
| Richest | 5844 | 18.3% |

Each of the 11 symptoms investigated were significantly (P<0.01) more likely to be reported by those testing positive as compared to those testing negative (Table 2). The most frequently reported symptoms among positive cases were headaches (58.0%), change in smell or taste (56.5%), fever (52.1%), cough (47.7%) and myalgia (44.1%). Table 2 also presents the prevalence ratios for each symptom and the 95% CI according to SARS-CoV-2 antibodies. The largest ratios between positive and negative subjects were observed for change in smell or taste (6.2-fold), fever (4.3-fold), shivering (3.3-fold) and myalgia (2.8-fold). The sensitivity and specificity for positive test results, for each symptom, are presented in Supplementary Table. The two symptoms with sensitivity above 50% and specificity above 85% were change in smell or taste, followed by fever.

**Table 2.** Prevalence of symptoms among subjects with positive and negative antibody tests for SARS-CoV-2, and prevalence ratios. The EPICOVID19 study, third wave.

| Symptom | Prevalence |  | Prevalence ratio | 95% CI |  |
| --- | --- | --- | --- | --- | --- |
|  | Positive | Negative |  | Lower bound | Upper bound |
| Headaches | 58.0 | 35.5 | 1.6 | 1.5 | 1.8 |
| Change in smell or taste | 56.5 | 9.1 | 6.2 | 5.6 | 6.8 |
| Fever | 52.1 | 12.2 | 4.3 | 3.9 | 4.7 |
| Cough | 47.7 | 22.2 | 2.1 | 1.9 | 2.4 |
| Myalgia | 44.1 | 15.7 | 2.8 | 2.5 | 3.1 |
| Sore throat | 33.8 | 16.6 | 2.0 | 1.8 | 2.3 |
| Diarrhea | 25.6 | 11.7 | 2.2 | 1.9 | 2.5 |
| Difficulty breathing | 23.1 | 9.4 | 2.5 | 2.1 | 2.8 |
| Shivering | 20.5 | 6.1 | 3.3 | 2.9 | 3.9 |
| Palpitation | 20.0 | 10.5 | 1.9 | 1.6 | 2.2 |
| Vomiting | 9.5 | 4.0 | 2.4 | 1.9 | 3.0 |

Of the 849 participants who tested positive for SARS-CoV-2 antibodies, only 12.1% (95%CI 10.1-14.5) reported none of the 11 symptoms and were therefore classified as asymptomatic, against 42.2% (95%CI 41.7-42.8) among those who tested negative (Figure 1). The mean (SE) number of symptoms for those who tested positive or negative were 3.91 (0.10) and 1.53 (0.01), respectively. Among those who tested positive, 63.5% had three or more symptoms, compared to 23.0% among those who tested negative.

**Figure 1.**
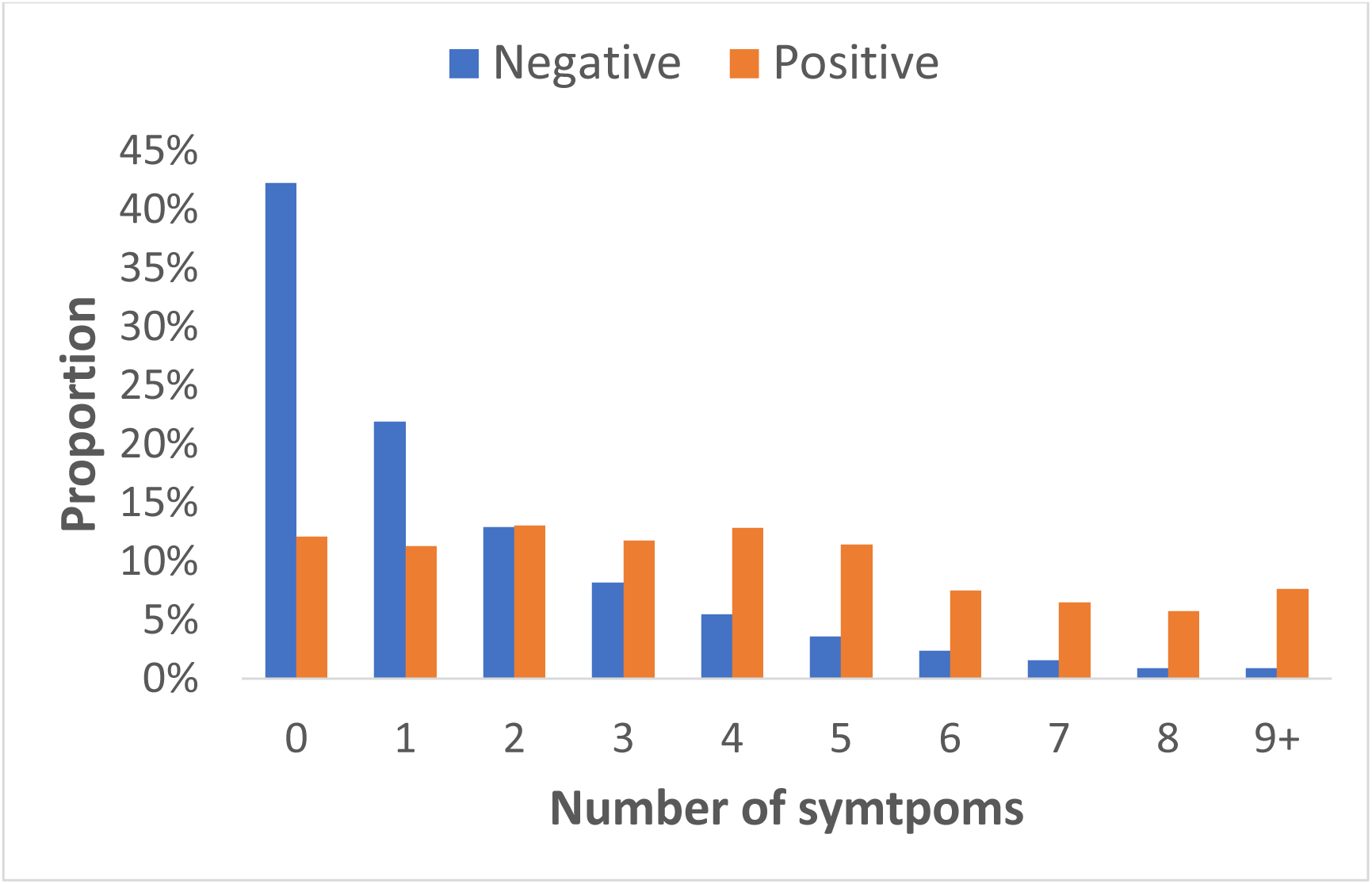
Distribution of the number of symptoms in individuals positive and negative for antibodies for SARS-CoV-2. The EPICOVID19 study, third wave.

In Figure 2 we present the mean number of symptoms among those who tested positive for SARS-CoV-2 as well as the prevalence of asymptomatic subjects, according to sociodemographic characteristics. The associations with ethnicity and household wealth were not significant. Symptoms were more frequent among women than men, and less frequent among individuals with primary or less schooling compared to those with secondary or higher education. The age distribution for the number of symptoms showed in inverse U-shaped pattern, with highest value at ages 30-39 years and the lowest means in children and adolescents. The corresponding figure for individuals without antibodies is included in the supplementary materials (Supplementary Figure 1).

**Figure 2.**
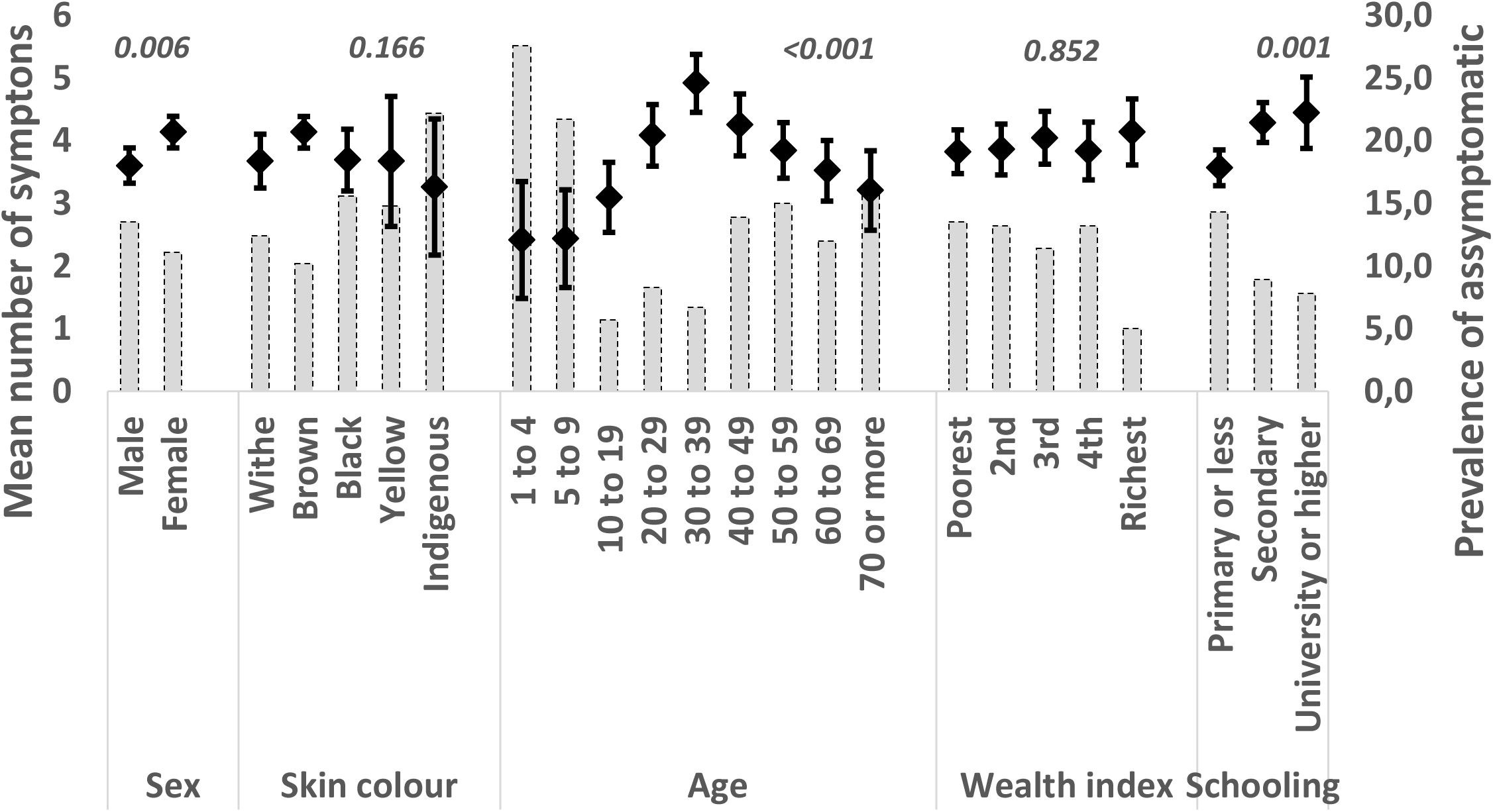
Mean number of symptoms and percent asymptomatic in subjects positive for antibodies against SARS-CoV-2, according to sociodemographic characteristics. The EPICOVID19 study, third wave. *Notes: diamonds represent the main Y axis for the mean number of symptoms (with their respective 95% CI). The bars represent the secondary Y axis for the prevalence of asymptomatic subjects*.

Figure 3 displays the results of the conditional inference tree analysis. Out of the 11 symptoms, this analysis selected three: change in smell or taste, fever and myalgia. Given the low overall seroprevalence, in all terminal nodes the prevalence was lower than 20%. Notably, the two thirds of the total sample who reported none of the three symptoms presented a markedly low seroprevalence of 0.8%, compared to 18.3% among those presenting fever, myalgia and change in smell or taste.

When an individual tested positive, we also tested other family members. Of the 90 positive subjects with at least one positive family member, 6.7% were asymptomatic, compared to 13.0% asymptomatic among 747 positive subjects without any positive family members (chi-squared = 2.97; P = 0.085). Lastly, we verified whether antibody prevalence levels in cities were associated with the frequency of symptoms among positive subjects, and found no such association (Supplementary Figure 2).

**Figure 3.**
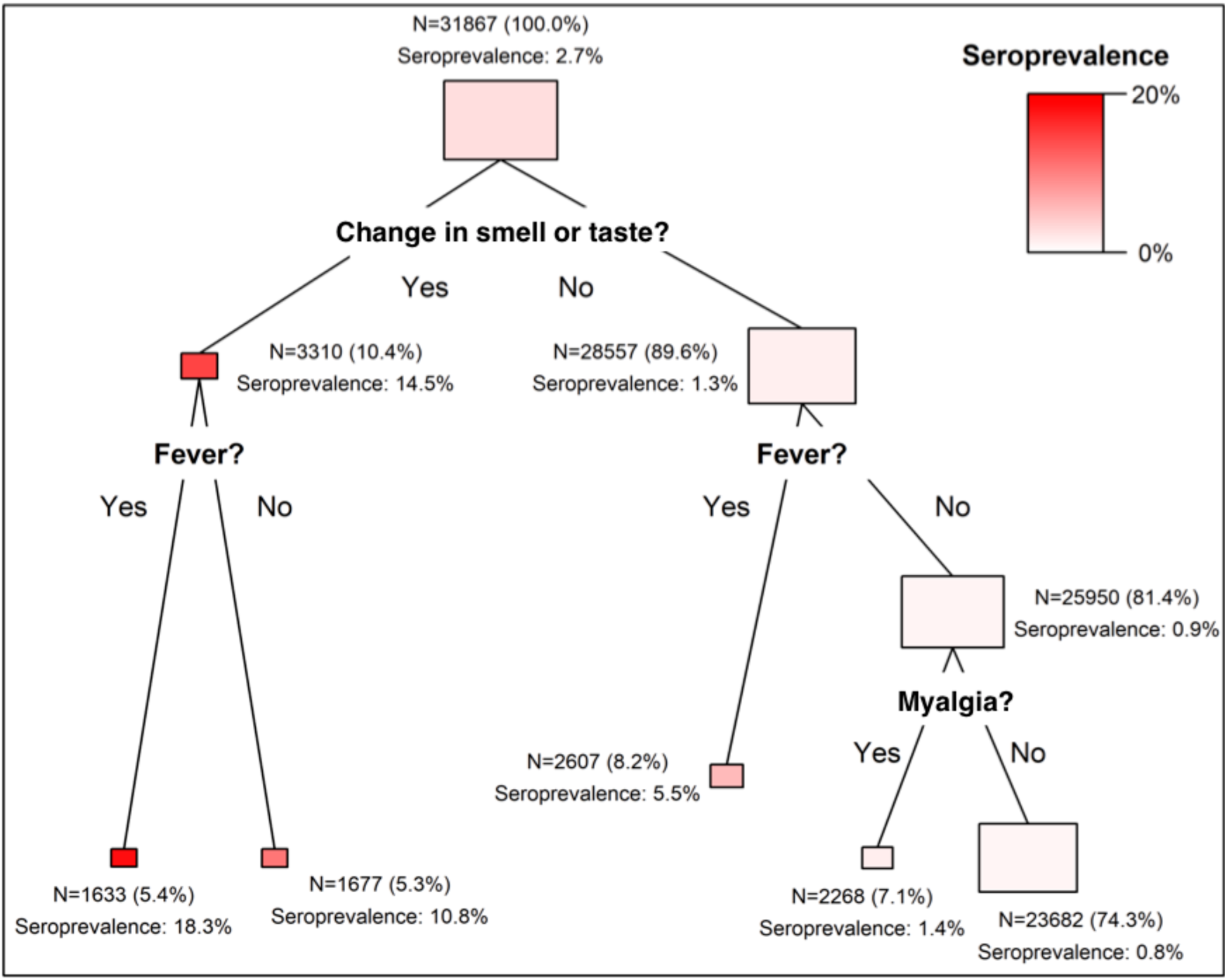
– Conditional inference tree of the association between symptoms (predictors) and seroprevalence for SARS-CoV-2. The EPICOVID19 study, third wave. The area of the rectangles corresponds to the proportion of the population contained in each node.

## DISCUSSION

In the first two waves of the EPICOVID19 nationwide survey, we identified that, contrary to what is often reported, most subjects with antibodies were symptomatic. However, symptoms had only been assessed for those with positive tests, and the information was collected after the individual had learned about the test result. We addressed the possibility of bias by asking all participants, regardless of the test result, in the third wave. The question on symptoms covered the four-month period since the first COVID-19 cases were reported in the country. The questionnaire was applied before the test result was known, so that respondents were blind to their serological status, and this allowed us to compare symptoms among those testing positive and those testing negative. Subjects with a previous diagnosis of COVID-19 and missing information for symptoms (0.73% of the whole sample) were excluded from the analyses in order to ensure that the respondents were not aware of their condition.

The above results from the third wave of the study confirmed a high prevalence of symptoms using a 4-month recall period; only 12.1% positive subjects were asymptomatic, compared to 42.2% of those without antibodies. Inclusion in our analyses of individuals who tested negative was useful for identifying which symptoms were most strongly associated with the presence of antibodies. For example, headaches were the most common symptom affecting 58.0% of those positive, but were also reported by 35.5% of those who tested negative, a prevalence ratio of only 1.6. In contrast, changes in smell or taste affected 56.5% among those who tested positive and 9.1% in the negative ones, respectively. This symptom provided the best discrimination, with a prevalence ratio of 6.2. Recent studies have shown that when SARS-CoV-2 enters the nasal and oral epithelium through the angiotensin-converting enzyme 2 (ACE2) and transmembrane serine protease 2 (TMPRSS2), it may cause damages to olfactory and gustatory receptor cells resulting in anosmia or ageusia^14, 15^.

Overall, symptoms were more frequent among females than males, in subjects aged 30-29 years and in those with higher education. Children and adolescents were substantially less likely to report symptoms than adults, which is compatible with the lower infection-fatality rates observed in these age groups^16^. In contrast, prevalence of symptoms fell with age from 30 to over 70 years, which does not reflect the age pattern in infection-fatality and case-fatality^17^. The difference in reported symptoms between women and men is also at odds with the higher case-fatality among males^18^.

Comparison of our findings on the prevalence of symptoms with the literature are affected by the settings in which studies were done, by the phase of infection, the duration of recall, and by the ways in which symptoms were recorded, as well as whether or not the subjects were aware or suspicious of being infected. The prevalence for asymptomatic subjects in the literature ranges from 4% to 75%^2-6, 19, 20^, whereas in our study it was 12.1%. We identified five published reviews that provided pooled prevalence estimates for symptoms^4, 5 21-23^ among individuals who tested positive in health facilities. We found lower prevalence (52.1%) for fever (pooled prevalence ranging from 78.4% to 92.8%) and cough (47.7% versus pooled prevalence ranging from 58.3% to 72.2%). Our estimates for myalgia (44.1%) and difficulty breathing (23.1%) were within the ranges reported in the studies (29.4% to 51.0%, and 20.6% to 45.6%, respectively). Lastly, prevalence of headache in our study (58%) was considerably higher than in the reviews (8.0% to 14.0%). One may assume the prevalence ranges of symptoms based on individuals who sought care in medical facilities would tend to be higher than in our population-based survey, but this was not the case, except for fever or cough.

Notably, change in smell or taste was not investigated in these review papers. We searched the literature and change in smell or taste or anosmia/ ageusia was identified in a multicenter European study with prevalence of 85.6% (anosmia) and 88.0% (ageusia)^24^ and a very low prevalence in a retrospective study in China (5.1% for hyposmia and 5.6% for hypogeusia)^25^, whereas we found 56.5%.

Besides the aforementioned symptoms, some studies have hypothesized that the angiotensin-converting enzyme 2 receptor (ACE2) is also expressed in the mucosa of the gastrointestinal (GI) tract and play lead to GI manifestations^26^. The pooled prevalence of GI symptoms has ranged in the literature from 7.4 to 12.5% for diarrhea (against 25.6% in our study), and 4.6% to 10.2% for nausea and/or vomiting (compared to 9.5% in our study)^26-28^.

It is likely that the information on symptoms from population-based studies, such as the one from Spain^29^, would be comparable to our study; however, the recall time in that study was two weeks, compared to up to four months in our survey. In this study, the only symptom specifically reported was anosmia, that was present around 27% of positive subjects, in the three waves.

The decision tree analyses were useful for identifying a subgroup of individuals who presented both fever and change in smell or taste, among whom seroprevalence was 18.3%, compared to only 0.8% among subjects that did not present these two symptoms, nor presented body aches.

It is clear from the literature that no single symptom correlates perfectly with SARS-CoV-2 infection, thus raising the possibility that the use of multiple symptoms might be appropriate for screening purposes. However, the literature on this topic is still scarce. A study using app-based self-reported data in the United States and in the United Kingdom identified that change in smell or taste is the single symptom most strongly correlated with infection and, using stepwise logistic regression, identified a prediction model that also includes fatigue, persistent cough and loss of appetite^30^. We also identified change in smell or taste as the single most predictive symptom, but the two additional symptoms prioritized in the conditional inference tree analysis were fever and myalgia. Given that the symptoms are partially correlated to one another, it is possible that models including different symptoms yield similar predictions, and would therefore be of similar practical use. Another app-based study including mostly individuals in the United Kingdom identified that, collectively, symptoms improve predicting prognosis^31^. This indicates that symptoms may be used not only for screening, but also for patient monitoring and planning health service needs.

Our study has limitations. Differentiation recall bias is a concern, particularly by using a 4-month recall period, but the alternative – as in the Spanish survey – was to ask for symptoms in a shorter, more recent period and potentially misclassifying individuals who had the disease in the past, and for whom antibodies remained detectable. In order to evaluate the likelihood of differential recall bias, we excluded the 242 participants who had a diagnosis of COVID-19 prior to the interview. Another limitation is the growing evidence that antibody levels decrease rapidly over time, for example by 14% in the same subjects in the Spanish study^29^, and in our own (unpublished) analyses comparing the first and third waves of the survey in cities with high initial prevalence. This would lead some individuals who had the disease to test negative, and yet report symptoms that occurred at the time of the episode. This type of bias would reduce the difference in reported symptoms among subjects who tested positive and negative. An additional limitation is the growing evidence that antibody levels decrease rapidly over time, for example by 14% in the same subjects in the Spanish study^28^, and in our own (unpublished) analyses comparing the first and third waves of the survey in cities with high initial prevalence. This would lead some individuals who had the disease to test negative, and yet report symptoms that occurred at the time of the episode. This characteristic of the dynamics of the infection would reduce the difference in reported symptoms among subjects who tested positive and negative.

Positive aspects of our study, on the other hand, included the population basis over an area of 8.5 million square km, the large sample size, collection of symptoms in positive and negative cases, and blinding of respondents as test results were only disclosed after the clinical history was collected.

In summary, our analyses show that most individuals with antibodies against SARS-CoV-2 report having presented symptoms, even though in most cases these were mild. Our findings can be used to implement surveillance systems in Brazil, which would help identify cases early and guide testing procedures.

## Data Availability

Data will become publicly available 30 days after completion of the fieldwork at http://www.epicovid19brasil.org/

## Acknowledgments

We acknowledge the support from Instituto Serrapilheira, Pastoral da Criança, the Brazilian Collective Health Association (ABRASCO) and JBS’s initiative ‘Fazer o Bem Faz Bem’.

Ana M B Menezes, Cesar G Victora, Fernando P Hartwig, Mariângela F Silveira, Bernardo L Horta, Aluísio J D Barros, Lúcia C Pellanda, Odir A Dellagostin, Claudio J Struchiner, Marcelo Burattini, Fernando C Barros and Pedro C Hallal contributed to the conception and design of the work, to the acquisition, analysis, and interpretation of data and the draft of the manuscript. Marilia A Mesenburg and Fernando C Whermeister contributed to the analysis of data. All authors have approved the submitted version and have agreed to be personally accountable for the author’s own contributions and to ensure that questions related to the accuracy or integrity of any part of the work, even ones in which the author was not personally involved, are appropriately investigated, resolved, and the resolution documented in the literature.

